# Willingness to Pay for Health Insurance in Sub-Saharan Africa: A Systematic Review Protocol

**DOI:** 10.1101/2025.05.16.25327764

**Authors:** Cyril E. Dzah, Theodora O.K. Amponsah, Richard A. Bonney, Daniel Opoku

## Abstract

**Introduction:** In sub-Saharan Africa, most people pay for healthcare directly out-of-pocket (OOP). This makes it harder for countries in the region to achieve Universal Health Coverage (UHC), as many people can’t afford care, resulting in poor health, and some people being pushed deeper into poverty due to the high cost of care. To reduce this burden and move closer to UHC, many African countries have introduced health insurance to raise funds for healthcare and reduce direct OOP spending. To make the health insurance more effective, it’s important to understand how willing people are to pay for health insurance, how much can be raised relative to GDP, and what factors influence their willingness to pay (WTP). This information can help shape health financing policies in the region.

**Methods and Analysis:** A systematic search will be conducted using PubMed, Google Scholar, Science Direct, relevant references from selected studies and grey literature that would be identified from the databases searched. The search will cover all available studies that focus on WTP for health insurance in sub-Saharan Africa from inception to 31^st^ May 2025. Key outcomes will include the percentage of people willing to pay, amount to be raised relative to GDP per capita, and the factors that frequently influence WTP. Each study will be carefully reviewed using the STROBE (strengthening the reporting of observational studies in Epidemiology) tool for quality, and the results will be combined and analysed using quantitative methods.

**Ethics and Dissemination:** Since this is a systematic review that does not involve recruiting or studying people directly, ethics approval is not required. The findings will be disseminated through conferences, symposia, and published in a peer-reviewed journal. They will also be shared with policymakers and the general public interested in health financing.

**PROSPERO registration number:** CRD420251027763

**STRENGTHS AND LIMITATIONS OF THIS STUDY:**

- Follows a structured and transparent systematic review methodology.
- Uses STROBE tool for critical appraisal of included studies.
- Includes standardization of amounts to be raised across countries using GDP per capita.
- Employs vote-counting to identify influencing factors.
- Limited to studies conducted in Sub-Saharan African countries published in English

## INTRODUCTION

In many Sub-Saharan African countries, access to healthcare often means paying out-of-pocket (OOP) [1,2]. For most people in the region, this can be a huge financial burden, making it difficult to access the care they need and pushing many into poverty [3–7]. This heavy reliance on OOP payments also makes it hard for countries in the region to reach the Universal Health Coverage (UHC) goal, where everyone can receive the needed quality healthcare without facing financial hardship [1,3,8,9].

To address the catastrophic and impoverishing effects of healthcare expenditures, governments must take decisive steps to protect people from high healthcare costs [4,5,7]. One way to do this is by collecting and managing enough money locally to support the health system, without depending too much on people’s direct payments at the point of care [5,8,10,11]. So far, many improvements in healthcare in sub-Saharan Africa have come from foreign aid. But this kind of support isn’t reliable or sustainable, as external funding has declined significantly with much of it directed to specific health problems and not the overall health system [3,6,11]. That’s why the focus is shifting toward raising money from within each country, through equitable, sustainable and effective public health financing strategies [6,10,12]. The United Nations and WHO have encouraged member countries to use prepayment systems, like health insurance, to reduce the need for cash or direct OOP payments at the point of care [5,8]. Even though some progress has been made in the area of health insurance across the region, many countries in the region are still far from achieving the UHC goal by 2030 [3,13].

Reaching UHC takes more than just setting up health insurance systems, it also requires making sure those systems can actually bring in enough money to support the healthcare needs of the population. Therefore, to move closer to the UHC goal and to protect people from financial risks related to healthcare, countries in sub-Saharan Africa need to prioritize health insurance mechanisms and one way is by understanding the health financing behaviour of the population. This study therefore systematically reviews evidence on the willingness to pay (WTP) for health insurance in the region, how much can be raised from the health insurance, and what factors influence the WTP. This information can help inform decision-making in health financing policies in the region.

## METHODS AND ANALYSIS

### Protocol Development

This study protocol follows the Preferred Reporting Items for Systematic Review and Meta-Analysis Protocols (PRISMA-P) guidelines. The protocol for this review is officially registered with the International Prospective Register of Systematic Reviews (PROSPERO) with registration number CRD420251027763

### Study Design

This is a systematic review of published primary studies examining WTP for health insurance in sub-Saharan Africa.

### Study Selection

The process for selecting studies for this review is shown in Figure 1. Two members of the research team will independently review each study to decide whether it should be included. First, they will screen the titles and abstracts. Studies that pass this initial stage will then be assessed in full. If there are any disagreements, the reviewers will discuss and come to a joint decision. For every study that is excluded, the reason for exclusion will be documented.

**Fig 1.**
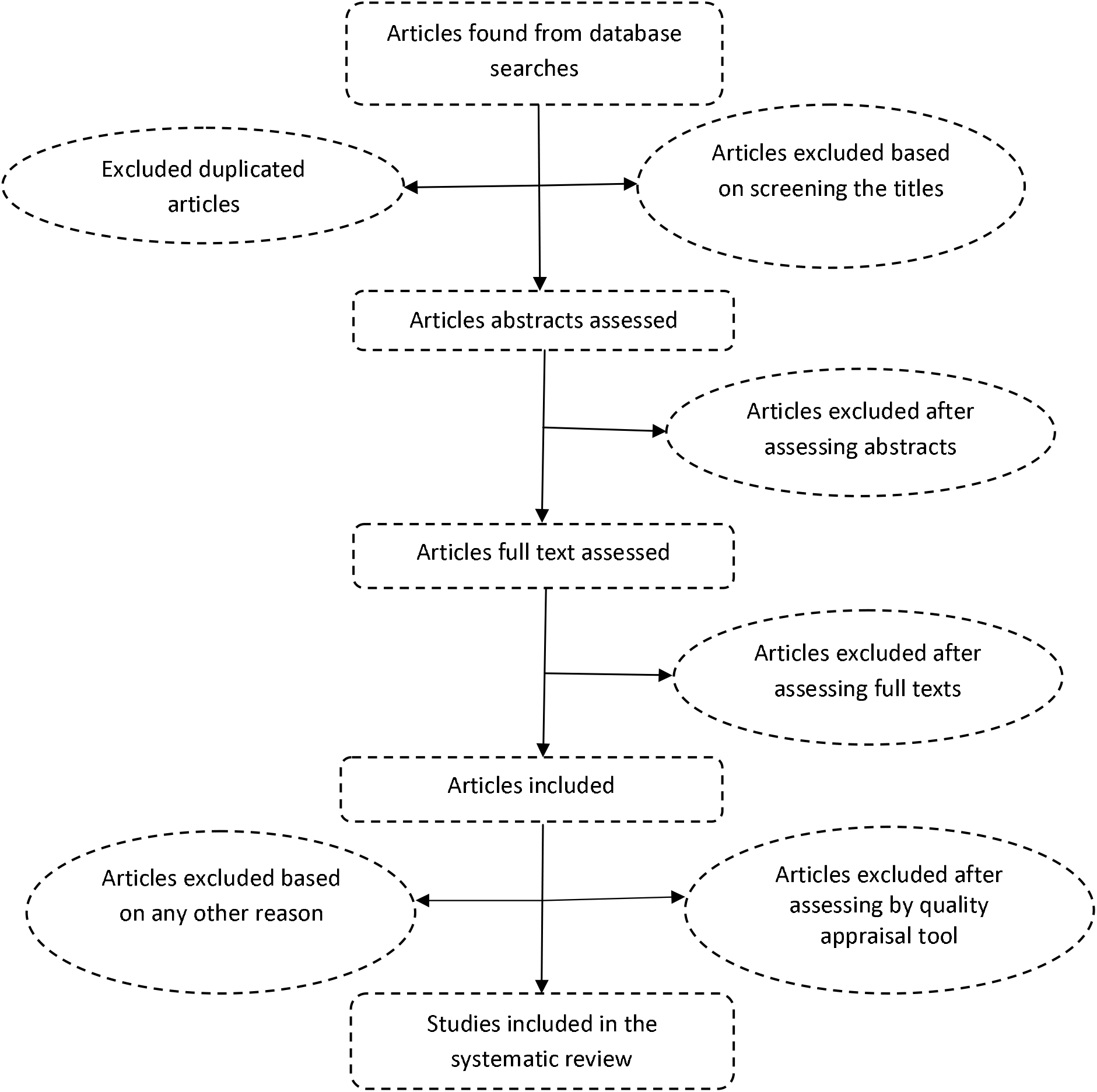
Article flowchart for studies of WTP for health insurance demonstrating how studies will be included in the systematic review.

### Eligibility Criteria

#### Inclusion Criteria

Primary studies that looked at households’ or individuals’ willingness to pay for health insurance in Sub-Saharan African countries will be considered for inclusion in the systematic literature review if they:

1. calculated the percentage of people willing to pay.
2. calculated an average amount of money the people were willing to pay or that will be raised.
3. collected data on factors that influence WTP.

### Exclusion Criteria

Studies will be excluded in this systematic review if they:

1. focused only on specific services or populations (e.g., maternal care, public servants).

### Information Sources and Search Strategy

Information for this review will be gathered through a comprehensive literature search covering all studies available up to May 31, 2025. The databases to be searched include PubMed, Google Scholar, Science Direct and grey literature identified through the searched databases. Additional relevant studies found in the reference lists of the included articles will also be considered. The search will be limited to studies documented in the English language.

Search terms were developed and combined using Boolean operators ‘AND’ and ‘OR’. The search term that would be used to search databases are as follows: “Willingness to pay” AND “health OR health insurance”. The complete PubMed search strategy developed is:

#1 Search willingness to pay * Field: Title/Abstract,

Limits: humans /English language

#2 Search health OR health insurance *Field: Title/Abstract,

Limits: humans / English language

Search #1 AND #2 Field: Title/Abstract, Limits: Humans / English language

### Data Extraction

Data will be extracted from all the included studies by two independent reviewers. The results will be compared for consistency. The information to be collected will include the study location, the contingent valuation method used, the year of study, the target population, the sample size, the variables that significantly affected WTP, and the amounts the participants are willing to pay.

### Quality Appraisal

The STROBE (strengthening the reporting of observational studies in Epidemiology) tool will be used to assess the quality of the studies included in this review. It ensures that each study clearly reports important aspects like its design, methods, and findings, helping to confirm that the evidence is reliable, transparent, and appropriate for analysis.

### Data Synthesis

Two approaches will be used for the analysis and synthesis of data:

i. Standardization of the various studies to ensure comparison and analysis of WTP amounts (the amount of money the people were willing to pay).
ii. Vote-counting method will be applied to identify the key variables that significantly influenced WTP.

### Standardization of the Various Studies

The standardization of the various studies will be done as follows:

- All included studies report the amount of money the participants were willing to pay.
- The amounts will be converted into effect size by calculating the annual amounts for studies that do not have annual amounts. The annual amounts will then be converted to US dollars using the exchange rate values reported in the various studies. For studies that did not report exchange rates, the International Monetary Fund (IMF) values will be used per the year the study was conducted.
- In the final stage, the amounts as a percentage of GDP per capita will be calculated per the GDP at the time the study was conducted.

### Descriptive Vote Counting

Vote-counting will be conducted using the following approach:

- The variables that influenced WTP for health in the included studies will be identified.
- The identified variables will be categorized into those with positive significant effects on WTP and those with negative significant effects on WTP as reported in the various included studies.
- A vote will be cast per a significant positive or negative effect of variables as reported and the total number of votes per variable documented.
- Variables that obtained three and above votes will be considered likely to influence WTP for health insurance.

## Data Availability

All data produced in the present work are contained in the manuscript

## Ethics and Dissemination

Ethical approval is not needed for this study, as it is a systematic review that does not involve recruiting or studying people directly. The findings will be shared at conferences and symposia, and communicated to the general public, policymakers, and stakeholders interested in health financing, and published in a peer-reviewed journal.

## Authors’ Contributions

Cyril E. Dzah (guarantor) developed the study protocol and wrote the initial draft of the manuscript. Daniel Opoku provided supervision throughout the development process. Richard A. Bonney, Theodora O. K. Amponsah and Daniel Opoku reviewed the protocol and provided feedback. All authors contributed to discussions on the study’s design and concept, and the final manuscript was reviewed and revised by all authors.

